# COVID-19 and human milk: SARS-CoV-2, antibodies, and neutralizing capacity

**DOI:** 10.1101/2020.09.16.20196071

**Authors:** Ryan M. Pace, Janet E. Williams, Kirsi M. Järvinen, Mandy B. Belfort, Christina D.W. Pace, Kimberly A. Lackey, Alexandra C. Gogel, Phuong Nguyen-Contant, Preshetha Kanagaiah, Theresa Fitzgerald, Rita Ferri, Bridget Young, Casey Rosen-Carole, Nichole Diaz, Courtney L. Meehan, Beatrice Caffe, Mark Y. Sangster, David Topham, Mark A. McGuire, Antti Seppo, Michelle K. McGuire

## Abstract

**Background:** It is not known whether SARS-CoV-2 can be transmitted from mother to infant during breastfeeding, and if so whether the benefits of breastfeeding outweigh this risk. This study was designed to evaluate 1) if SARS-CoV-2 RNA can be detected in milk and on the breast of infected women, 2) concentrations of milk-borne anti-SARS-CoV-2 antibodies, and 3) the capacity of milk to neutralize SARS-CoV-2 infectivity.

**Methods:** We collected 37 milk samples and 70 breast swabs (before and after breast washing) from 18 women recently diagnosed with COVID-19. Samples were analyzed for SARS-CoV-2 RNA using RT-qPCR. Milk was also analyzed for IgA and IgG specific for the nucleocapsid protein, receptor binding domain (RBD), S2 subunit of the spike protein of SARS-CoV-2, as well as 2 seasonal coronaviruses using ELISA; and for its ability to neutralize SARS-CoV-2.

**Results:** We did not detect SARS-CoV-2 RNA in any milk sample. In contrast, SARS-CoV-2 RNA was detected on several breast swabs, although only one was considered conclusive. All milk contained SARS-CoV-2-specific IgA and IgG, and levels of anti-RBD IgA correlated with SARS-CoV-2 neutralization. Strong correlations between levels of IgA and IgG to SARS-CoV-2 and seasonal coronaviruses were noted.

**Conclusions:** Our data do not support maternal-to-child transmission of SARS-CoV-2 via milk; however, risk of transmission via breast skin should be further evaluated. Importantly, milk produced by infected mothers is a source of anti-SARS-CoV-2 IgA and IgG and neutralizes SARS-CoV-2 activity. These results support recommendations to continue breastfeeding during mild-to-moderate maternal COVID-19 illness.

## Introduction

The global spread of severe acute respiratory virus 2 (SARS-CoV-2), the causative agent of coronavirus disease 2019 (COVID-19), has led to concerns over mother-to-child transmission, including via breastfeeding. Several studies have reported the presence of SARS-CoV-2 RNA in human milk,^1-4^ whereas others have not^5-9^ (Table S1). Most previous studies are limited because they followed only a few participants, were cross-sectional, and/or failed to report how milk was collected and/or analyzed. Thus, considerable uncertainty remains regarding whether human milk is capable of transmitting SARS-CoV-2 from mother to infant.

This paucity of rigorous methodology combined with inconsistency of viral RNA detection across studies has led to conflicting and changing recommendations regarding temporary separation of infants from mothers with COVID-19 and regarding whether infants should nurse directly at the breast or receive expressed milk from a bottle.^10-13^ Alongside the uncertainty about the risks of breastfeeding in the context of maternal COVID-19, it is well established that breastfeeding reduces the risk of myriad short- and long-term infectious and noninfectious conditions.^14^ Further, even a short delay in initiation of breastfeeding can interfere with the establishment of lactation^15^ and increase risks of infant morbidity and mortality.^16-18^

Many of the health-promoting effects of breastfeeding are due to the provision of passive immunity via immunoglobulins and other bioactive factors (e.g., lactoferrin), and previous studies have shown that milk-borne antibodies are produced in response to viral infection.^19-22^ However, few studies have examined the presence of antibodies to SARS-CoV-2 in human milk.^23,24^ In one recent study, milk from 12 of 15 women previously infected with SARS-CoV-2 contained IgA that was reactive to the receptor binding domain (RBD) of the SARS-CoV-2 spike protein.^24^ They also reported that antibodies in milk from previously infected women and milk collected prior to December 2019 (prepandemic) exhibited low-level cross-reactivity to RBD. However, levels of secretory IgA with reactivity to RBD were higher in milk from previously infected women. Cross-reactivity of antibodies in serum samples collected from healthy individuals and those infected with seasonal human non-SARS coronaviruses (sCoV) have also been reported.^25^ This cross-reactivity is thought to stem from homology of the spike protein of sCoVs and SARS-CoV-2. The extent to which milk-borne antibodies have cross-reactivity to sCoV and whether these cross-reactive antibodies are associated with neutralization of SARS-CoV-2 is currently not known.^26^

The primary objective of this study was to determine whether SARS-CoV-2 can be detected in milk produced by, and on the breast skin of, women recently diagnosed with COVID-19 utilizing rigorous collection and analytical techniques. We also aimed to quantify anti-SARS-CoV-2 IgA and IgG in milk and the capacity of milk to neutralize SARS-CoV-2. Because subclinical mastitis has been associated with higher viral loads in milk^27^, we also documented sodium-to-potassium ratios (Na/K) in milk, a biomarker of subclinical mastitis.

## Methods

### Experimental design and clinical data collection

This prospective study was carried out using a repeated-measures, longitudinal design. To be eligible, women needed to be ≥18 years of age, lactating, and have received a positive test result for COVID-19 in the previous 8 days. Subjects were recruited through social media; word-of-mouth; and assistance of national maternal and child health organizations and local hospitals. All participants gave informed consent, and procedures were approved by the Institutional Review Boards at the University of Idaho (20-056, 20-060), the University of Rochester Medical Center (1507), and Brigham and Women’s Hospital (2020P000804). Surveys were administered by telephone to ascertain timing of maternal/infant COVID-19 symptoms, reproductive history, breast health, breastfeeding practices, demographics, and anthropometrics.

### Milk and breast swabs

All collection kits were assembled aseptically by study personnel wearing masks and gloves and were individually packaged to reduce potential contamination. Mothers were instructed in clean techniques to obtain samples, including use of gloves and masks. Milk and swabs of the nipple/areola (“breast swabs”) were either self-collected in participants’ homes (with virtual assistance provided by study personnel), or at a hospital (participants Q and R were admitted to the postpartum unit at the time of sample collection). Breast swabs were collected before and after washing the breast with soap and water and prior to milk collection. Women collected up to 30 mL of milk using the provided sterile manual breast pump (Harmony, Medela) and sterile collection containers. Details regarding sample collection are provided in the Supplemental Appendix.

Following collection, samples were immediately frozen in the subject’s freezer until shipped in a cooler containing frozen cold packs to the University of Idaho (UI) or University of Rochester Medical Center (URMC). Samples collected from subjects Q and R were frozen at - 80°C and shipped on dry ice to UI. Once received, samples were processed immediately for RNA extraction and/or stored at -80°C until further analysis. As needed, samples were shipped on dry ice between UI and URMC. Milk samples collected from 10 healthy women located in the Rochester, NY area for general assay development purposes prior to December 2019 were used as prepandemic control samples.

### SARS-CoV-2 RNA

Details related to RNA extraction and assay validation are provided in the Supplemental Appendix. Briefly, RNA was extracted from milk (both at UI and URMC), breast swabs (UI), and extraction controls (both at UI and URMC) using the Quick-DNA/RNA Viral MagBead kit (Zymo Research, Irvine, CA) with addition of the DNase I treatment on extracted nucleic acids following the manufacturer’s protocol. Detection of SARS-CoV-2 viral RNA in milk was independently determined in both UI and URMC laboratories using the CDC-designed 2019-nCoV RT-qPCR assay^28^, validated in both laboratories for use with human milk. Per the CDC protocol, samples with Ct values <40 were considered positive.

### Sodium and potassium

For milk samples with sufficient volume (33 of 37), sodium (Na) and potassium (K) concentrations were quantified in 200 µL of milk using LAQUAtwin ion selective meters (Na-11 and K-11, respectively; Horiba Ltd., Kyoto). Prior to use, each meter was conditioned and calibrated according to vender specifications. After each measurement, meters were rinsed with Nanopure water, wiped dry, and allowed to reach zero. A Na to K ratio (Na/K) >0.6 was interpreted to indicate subclinical mastitis.^29,3^

### Antibodies

Concentrations of milk-borne IgA and IgG reactive to the SARS-CoV-2 spike (both S2 subunit and RBD) and nucleocapsid (N) proteins and spike proteins of seasonal coronaviruses 229E and OC43 were measured in milk samples by ELISA as previously described.^31^ Briefly, Nunc MaxiSorp 96-well plates (Thermo Fisher, Waltham, MA) were coated with optimized concentrations of antigens (1-5 μg/mL) overnight at 4°C. Coated plates were blocked for 1 h before the addition of serial 2-fold dilutions of samples. After a 2 h incubation at room temperature, plates were washed and bound IgG and IgA were detected with alkaline phosphatase-conjugated anti-human IgG (clone MT78; Mabtech, Stockholm, Sweden) and anti-human IgA. Bound antigen-specific antibodies were detected by adding p-nitrophenyl phosphate substrate (Thermo Fisher). Absorbance was read at 405 nm after color development. A weight-based concentration method was used to assign antigen-specific antibody titers in test samples.^31,32^

### SARS-CoV-2 neutralization

The neutralizing activity of milk against SARS-CoV-2 was measured by microneutralization (MN) assay. Briefly, duplicates of delipidated milk were serially diluted 2-fold in virus diluent and incubated with 100 TCID50 of SARS-CoV-2 virus (Hong Kong/VM20001061/2020 isolate) in 96-well flat-bottomed plates for 1□h at 37°C. After incubation, Vero E6/TMPRSS2 cells (kindly provided by Yoshihiro Kawaoka, National Institute of Infectious Diseases, Japan; 25,000 cells/well)^33^ were added to the virus/sample mixtures. Plates were incubated for 48 h at 37°C, when a cytopathic effect was evident in virus-only control wells. Cells were then fixed with 6% paraformaldehyde for 30 min and washed and stained with crystal violet for 1 h. The MN titer was identified as the highest dilution of sample that showed 50% neutralization based on the appearance of the stained cell monolayer as compared with the virus control well.

### Statistical analysis

Except where noted, all statistical analyses were performed using R (version 3.6.1). Statistical testing of antibody concentrations and MN titers were performed using parametric tests on log-transformed data. Linear regression was performed using either lm() or rlm() function in R as appropriate.

## Results

### Participants and samples collected

Eighteen women with a recent diagnosis of laboratory-confirmed COVID-19 participated in the study. On average, women were 34.2 ± 4.7 yr old and 6.8 ± 7.8 mo postpartum. Additional characteristics of study participants and their infants are presented in Table 1. Of the 18 participants, all but three had symptoms related to COVID-19, with the most common being loss of smell/taste (11/18), headache (10/18), and fatigue (7/18) (Fig. 1A). Two of the three participants who were asymptomatic throughout the course of the study were initially tested for routine surveillance prior to being admitted to the hospital for labor and delivery (participants L and M); while the other (participant N) was tested due to a potential occupational exposure. Two of six infants tested for COVID-19 had a positive result.

**Table 1.** Study participant characteristics (*n* = 18). Categorical data are given as number of participants and, in parentheses, percent of total. Continuous data are provided as means ± standard deviations. *Definitions put forth by the US Centers for Disease Control and Prevention were used for BMI categories. **Missing data from 1 individual.

| Maternal Characteristics (*n* = 18) |  | Infant Characteristics (*n* = 18) |  |
| --- | --- | --- | --- |
| Age (y) | 34.2 ± 4.7 | Female | 9 (50) |
| Race/Ethnicity |  | Gestational Age (wk) | 38.6 ± 1.7 |
| *Black, Hispanic* | 1 (6) | Birth Weight (g) | 3372 ± 560 |
| *Black, Non-Hispanic* | 1 (6) | Birth Length (cm)** | 50.3 ± 2.7 |
| *Pacific Islander* | 1 (6) | Breastfeeding Status |  |
| *White, Hispanic* | 1 (6) | *Exclusive* | 5 (28) |
| *White, Non-Hispanic* | 14 (78) | *Mixed Feeding* | 13 (72) |
| Body Mass Index (kg/m<sup>2</sup>)* | 28.9 ± 4.8 | COVID-19 Diagnostic Test | 6 (33) |
| *Normal/Healthy* | 5 (28) | *Negative Result* | 4 (67) |
| *Overweight* | 7 (39) |  |  |
| *Obese* | 6 (33) |  |  |
| Parity (#) | 1.9 ± 1.1 |  |  |
| Cesarean Delivery | 6 (33) |  |  |
| Time Postpartum (mo) | 6.8 ± 7.8 |  |  |
| History of Mastitis** | 1 (7) |  |  |
| Symptomatic at or Prior to Enrollment | 14 (78) |  |  |
| Developed Symptoms after Enrollment | 1 (6) |  |  |

**Figure 1.**
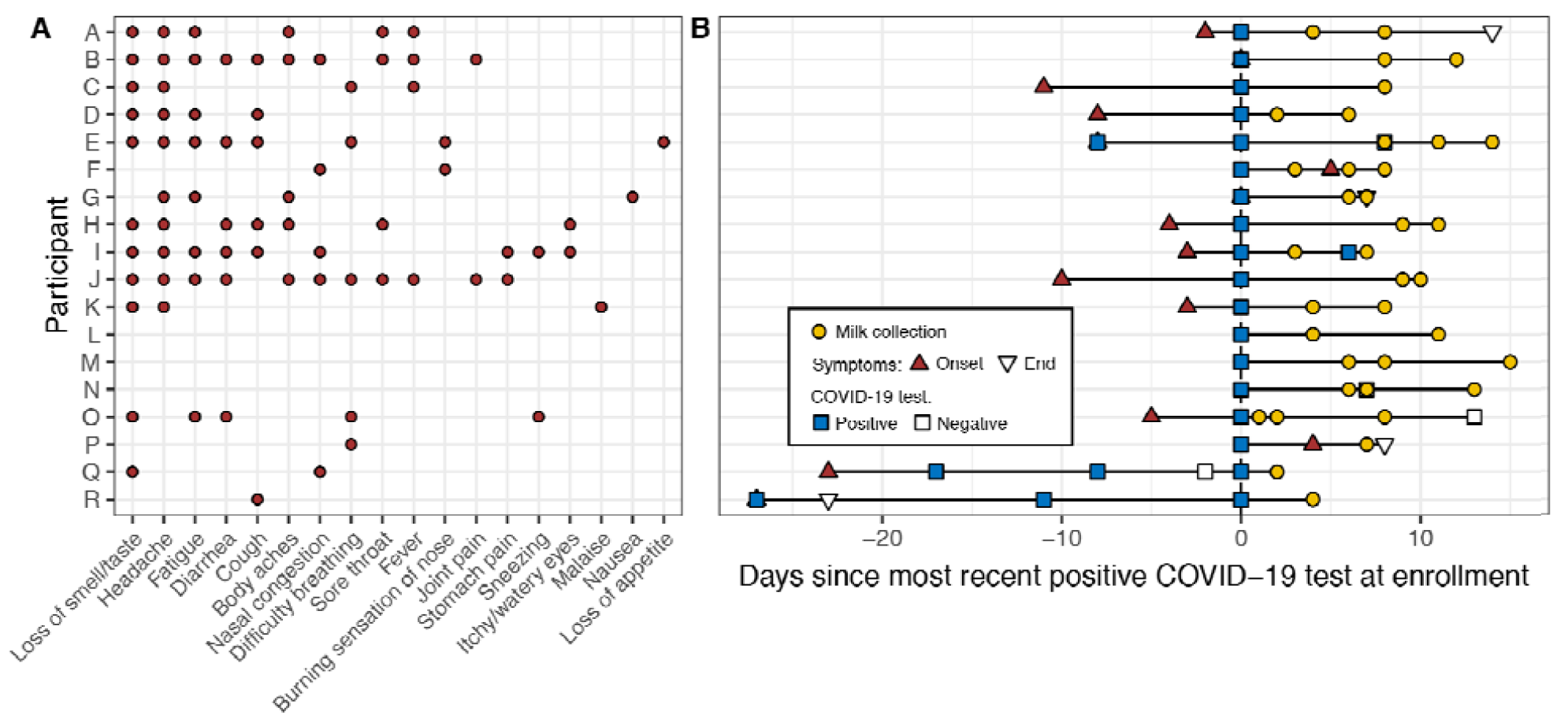
Overview of participants’ (A) COVID-19 symptoms, and (B) COVID-19 diagnostic testing and sampling. In panel B, the time “0” represents the most recent positive COVID-19 test at enrollment. Participants E, I, N, O, Q, and R had additional COVID-19 tests performed; all were positive except for N’s and O’s second test and Q’s third test which were negative (open squares). Participants L, M, and N were asymptomatic during the study period, and participants F and P developed symptoms after first testing positive.

We collected and analyzed 37 milk samples (Fig. 1B). Repeated samples were collected from 14 participants. Among women with symptoms at enrollment or who developed symptoms during the study, 6 provided samples within the first week of symptom(s), with the earliest sample collected 2 d prior to symptom(s) onset. This participant was initially tested for COVID-19 because of a close family exposure even though she was not currently symptomatic. Across all participants, the first sample was collected on average 12.0 ± 8.9 d after symptom onset. Breast swabs were collected from 15 women, although participant F collected swabs prior to breast cleaning and then after milk collection (rather than before milk collection) (Table S2).

### SARS-CoV-2 RNA and Na/K in milk

None of the milk contained detectable SARS-CoV-2 RNA. RT-qPCR findings were not modified by the milk fraction tested (i.e., whole milk or supernatant), and results were concordant between laboratories. Milk Na/K ratios ranged from 0.2 to 10.9 (0.5, median) with 12 (36%) samples having an elevated ratio (>0.6), suggesting subclinical mastitis in 9 participants.

### SARS-CoV-2 RNA on breast swabs

Of the 70 swabs tested, eight had evidence of SARS-CoV-2 RNA (Table S2). One swab collected prior to breast washing tested conclusively positive with Ct values <40 in both duplicates for both the N1 and N2 targets. Two additional swabs collected prior to breast washing had detectable signal in both duplicates for one of the SARS-CoV-2 targets, but only one duplicate for the other target. Five swabs had detectable signal in just one duplicate for one target.

### Anti-SARS-CoV-2 IgA and IgG in milk

Concentrations of anti-SARS-CoV-2 IgA were higher than those of IgG (Fig. 2A). Milk produced by women with COVID-19 had higher anti-RBD IgA and IgG concentrations than milk collected from women before the pandemic (*p*=0.000015 and p=0.00098, respectively). This pattern was also evident for anti-S2 and anti-N IgG (*p*=0.0006 and *p*=0.000089, respectively), but not IgA. Aside from the fact that prepandemic milk contained higher levels of IgA to sCoV 229E than milk produced during the pandemic (*p*=0.054), there was little difference between milk collected from study participants and prepandemic samples in terms of milk IgA and IgG to the full-length S proteins of sCoV 229E and OC43. Concentrations of IgA to SARS-CoV-2 and sCoV were correlated, particularly in milk produced by women with COVID-19 (Fig. 2B).

**Figure 2.**
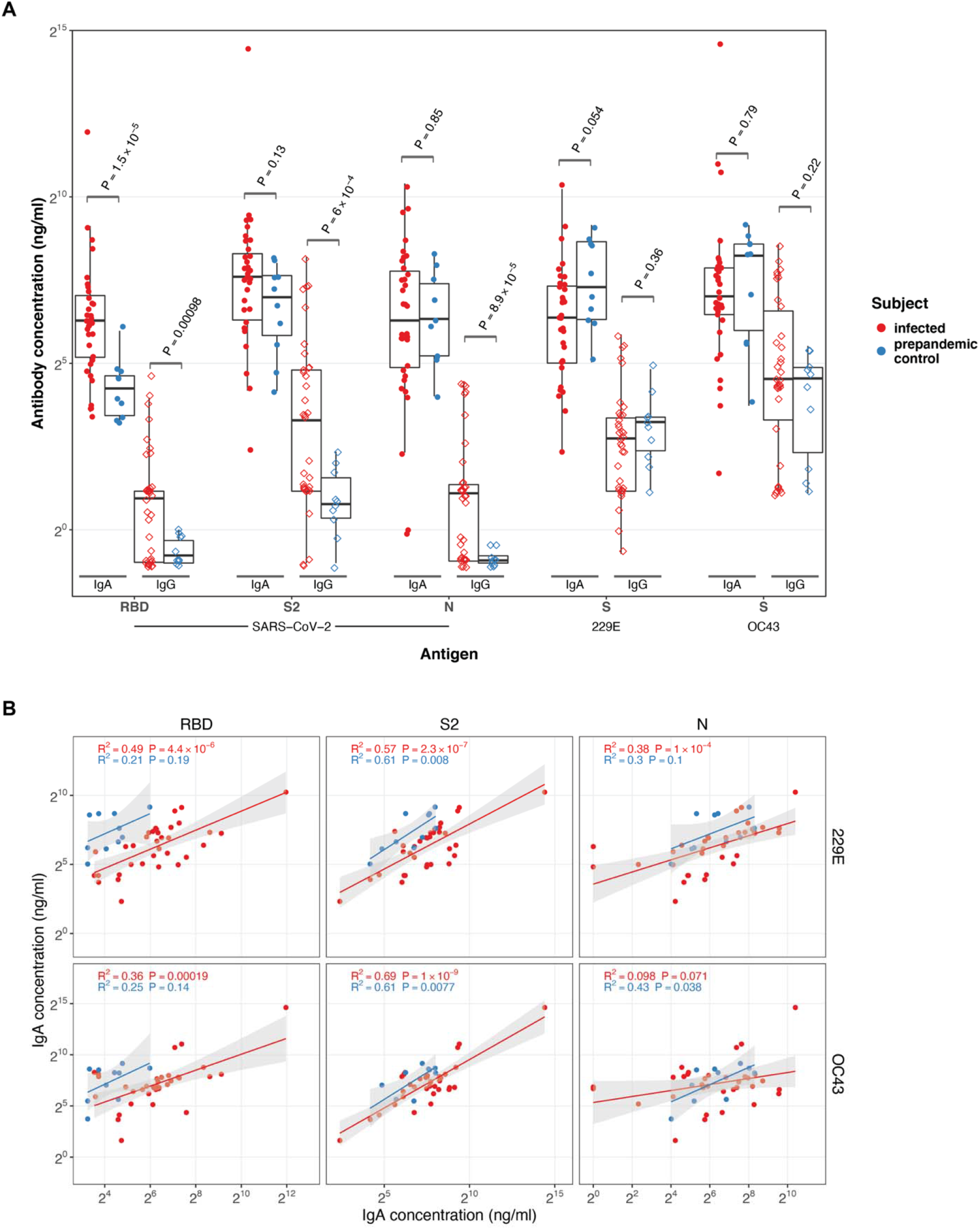
Milk antibody concentrations. Panel A shows IgA (filled circles) and IgG (open diamonds) to coronavirus antigens in milk produced by COVID-19 (red) infected and healthy, prepandemic (blue) women. Antibody concentrations were measured using ELISA specific to the RBD and S2 domains of the spike and nucleocapsid (N) proteins of SARS-CoV-2, and S proteins from human coronaviruses 229E and OC43. Panel B shows correlations between IgA concentrations to SARS-CoV-2 antigens RBD, S2, and N and IgA concentrations to S proteins from 229E and OC43 in milk produced by COVID-19 infected (red) and healthy, prepandemic (blue) women. A linear model was fit to log transformed IgA concentrations to give r-squared and p-values as indicated. Model prediction (red or blue line as above) is shown with the 95% confidence interval (grey shading).

### Neutralizing capacity of milk

A total of 21 of 34 milk samples (62%) collected from women with COVID-19 were found to neutralize SARS-CoV-2 infectivity *in vitro*. Although neutralization titers correlated with concentrations of IgA to all SARS-CoV-2 antigens tested (Fig. 3A and Fig. S1), in a regression model that included all antigen targets, only the SARS-CoV-2 RBD had a significant β (*p*=0.0125), consistent with neutralization primarily by anti-RBD antibodies. Furthermore, an analysis of sequential milk samples collected from study participants identified increases in anti-RBD IgA concentrations that were associated with elevated MN titers (Fig. 3B).

**Figure 3.**
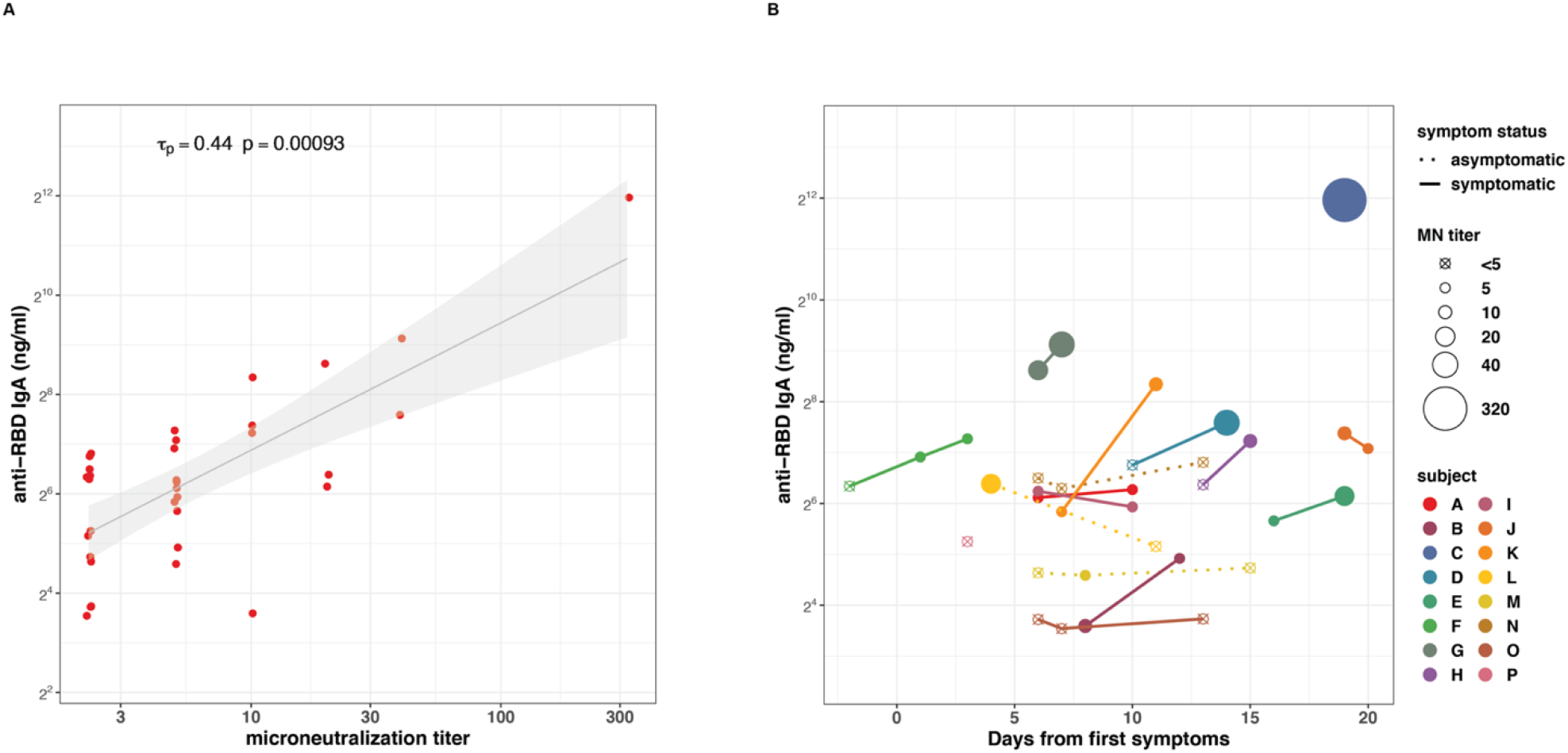
Correlation of milk IgA concentrations with microneutralization titers. Panel A shows correlation between concentrations of IgA specific to SARS-CoV-2 RBD and microneutralization titers in milk produced by COVID-19 infected women. Kendall rank correlation T_p_ and associated p-value as shown. A linear model was fit to log transformed IgA concentrations and MN titers and is shown (black line) with 95% confidence interval (grey shading) for visualization purposes only. Panel B shows anti-RBD IgA concentrations and microneutralization titers over time. Each color and corresponding point or set of connected points represent one participant. Time of collection is indicated on x-axis and antibody concentration on y-axis. MN titer of each sample is reflected in the size of each point. Asymptomatic participants are indicated using a dotted line connecting points.

## Discussion

Although human milk is considered the best source of nutrition for most infants, the onset of the global COVID-19 pandemic and our lack of understanding on SARS-CoV-2 transmission has caused confusion around whether infected mothers should be temporarily separated from their infants, as well as whether breastfeeding should be initiated and/or continued during maternal COVID-19 illness. In this prospective study we collected milk and breast swabs from women with COVID-19 and tested them for the presence of SARS-CoV-2 RNA. We also analyzed the milk for IgA and IgG targeting SARS-CoV-2 and the ability of the samples to neutralize SARS-CoV-2 infectivity.

Using methods validated and replicated across two laboratories, and consistent with most previous reports, we did not detect SARS-CoV-2 RNA in any of the milk samples. Although a single breast swab collected from the breast before it was cleaned was found conclusively to contain SARS-CoV-2 RNA, a swab collected after the breast was washed did not. It is noteworthy that several of the women in our study had evidence of subclinical mastitis, which has previously been shown to be positively related to milk RNA viral load in HIV-infected women.^27^ Nonetheless, we detected no SARS-CoV-2 in their milk. Together, our results suggest that milk may not act as a vehicle for mother-to-child transmission of SARS-CoV-2, although viral exposure via breast skin is possible. Our lack of detection of viral RNA on the breast after washing supports existing recommendations for women to take precautions during breastfeeding and/or expression of milk (e.g., practicing respiratory and hand hygiene, cleaning pump parts prior to and after use) to reduce the potential for viral transmission.

Importantly, we detected anti-SARS-CoV-2 antibodies in milk, primarily IgA but also IgG, albeit at lower concentrations than those reported for serum of actively infected patients.^34^ Concentrations of anti-SARS-CoV-S2 antibodies correlated strongly with those of the other tested sCoV spike proteins in milk produced by study participants and those produced prior to the pandemic. This pattern of cross-reactivity may reflect structural similarity among the proteins, and likely reflects a recall response from prior exposure to sCoV. However, generation of RBD-reactive antibodies likely requires generation of new B-cell populations, because the RBD of the SARS-CoV-2 spike protein shares little sequence homology with other SCoVs.

While the detection of SARS-CoV-2 RNA in milk and/or on breast is of concern, it does not necessarily indicate presence of viable virus. In the only study that has assessed viability of SARS-CoV-2 in milk, a single milk sample positive for SARS-CoV-2 RNA did not contain replication-competent virus^35^. Unfortunately, in our study we were unable to determine viability of SARS-CoV-2 in any of the breast swabs positive for SARS-CoV-2 RNA because the entire sample was needed for RNA detection. Future studies should determine the viability of any SARS-CoV-2 found in milk and/or the breast.

Our study has several strengths, including the use of rigorous collection methods; close temporal proximity of sample collections to COVID-19 diagnosis; validation of analytical methods for human milk; replication of RT-qPCR analyses across laboratories; and analysis of both risks and benefits of milk constituents. We also acknowledge that this study has limitations. For instance, most samples were collected from women after onset of symptoms, limiting generalizability to pre-symptomatic women. Additionally, none of our participants was hospitalized due to COVID-19. As disease severity may be related to viral titer^38^, it is possible that milk produced by individuals with more severe COVID-19 could contain SARS-CoV-2. The short duration of the follow-up period also does not allow characterization of the durability of the milk IgA and IgG responses. Initial reports on serum IgG response may suggest a relatively short-lived response^36,37^, and no data exist on the presence of long-lived memory B-cells in context of SARS-CoV-2. Milk IgA, representing a mucosal response, may have its own pattern of durability.

In summary, we did not detect SARS-CoV-2 RNA in milk collected from women with mild-to-moderate COVID-19. However, we demonstrated that milk contains anti-SARS-CoV-2 antibodies and that their concentrations are correlated with milk’s ability to effectively neutralize SARS-CoV-2 infectivity. We found evidence of SARS-CoV-2 on the breasts of several women, but it is unclear whether this RNA reflects viable virus. Taken together with the well-documented benefits of breastfeeding to maternal and infant health, our data support recommendations to encourage breastfeeding in women with mild-to-moderate COVID-19 illness.

## Data Availability

All requests for data and materials should be addressed to the corresponding authors.

## Acknowledgements

This study was funded by a grant from the Bill and Melinda Gates Foundation, in-kind support from Medela and Milk Stork, US National Institutes of Health (R01 HD092297-03, U01 AI131344-04S1), Washington State University Health Equity Research Center, and the University of Idaho Agricultural Experiment Station.

## Notes

### Competing Interest Statement

The authors have declared no competing interest.

### Author Declarations

All participants gave informed consent, and procedures were approved by the Institutional Review Boards at the University of Idaho (20-056, 20-060), the University of Rochester Medical Center (1507), and Brigham and Women's Hospital (2020P000804).

